# Association between female genital schistosomiasis and high-risk human papillomavirus among women of reproductive age in Zambia: the Schista study

**DOI:** 10.1101/2025.11.27.25341145

**Authors:** Olimpia Lamberti, Rhoda Ndubani, Jennifer Fitzpatrick, Emily Webb, Nkatya Kasese, Beatrice Nyondo, Barry Kosloff, Maina Cheeba, Philippe Mayaud, Morgan E. Lemin, Pytsje T. Hoekstra, Govert Van Dam, Paul Corstjens, Lisette Van Lieshout, Bonnie Webster, Helen Ayles, Isaiah Hansingo, Bodo Randrianasolo, Dingase Kuwemba, Bellington Vwalika, Paul Kamfwa, Kwame Shanaube, Helen Kelly, Amaya L. Bustinduy

## Abstract

**Background:** Female genital schistosomiasis (FGS) is a chronic gynaecological disease caused by the deposition of *Schistosoma haematobium* eggs in the female genital tract. It is highly prevalent in sub-Saharan Africa (SSA), the region with the highest cervical cancer incidence and mortality rates globally. Persistent infection with high risk (HR-) human papilloma virus (HPV) is necessary for cervical cancer development. We aimed to determine the cross-sectional association between FGS and HR-HPV genotypes in women across three communities in Zambia.

**Methods:** Women aged 15-50, sexually active, not menstruating or pregnant, were recruited at home by community health workers. Participants provided two cervicovaginal self-swabs, a urine sample, collected HIV and *Trichomonas vaginalis* self-tests and completed a questionnaire. At clinic follow-up, midwives collected two cervicovaginal swabs and cervical images with point-of-care colposcopy (EVA System, MobileODT^iZI^). Swabs were analysed for 14 HR-HPV types (GeneXpert^iZI^) and for *Schistosoma* DNA (ITS-2 qPCR). Urine samples were analysed for *Schistosoma* ova by microscopy and for circulating anodic antigen (CAA) to detect active infection. *Visual FGS* was defined as colposcopic identification of specific genital lesions. *Molecular FGS* was defined as *Schistosoma* qPCR positivecervicovaginal swabs.

**Results:** A total of 2,532 women (median age 28 years [IQR:22-36]) were recruited at home and 67% (1,694/2,532) completed clinic follow-up. Prevalence of *visual FGS, molecular FGS,* and HR-HPV were 35.2% [595/1,691], 6.5% [165/2,532], and 28.7% [690/2,401], respectively. HIV seropositivity was 17.5% (443/2,530), with 91.2% (406/443) self-reporting the use of antiretroviral therapy (ART). Risk factors crudely associated with HR-HPV infection included younger age, marital status, higher number of pregnancies, *molecular FGS* positivity, and urinary *S. haematobium* positivity detected using microscopy. There was evidence of a weak association between *molecular FGS* and all HR-HPV (adjusted Odds Ratio [aOR]=1.3, 95% Confidence Intervals [CI] 0.9-1.9). Women with *molecular FGS* were significantly more likely to test positive for HR-HPV 16/18/45 (aOR=1.7, 95%CI 1.0-2.8). No significant association was observed between *visual FGS* and HR-HPV infection (crude [c] OR = 0.9, 95% CI 0.7-1.1).

**Conclusions:** To our knowledge, this is the first study to jointly screen for FGS and HR-HPV in Zambia and to report an association between the most oncogenic HR-HPV types and *molecular FGS*. These findings call for the need for integrating FGS and HR-HPV screening strategies to address the dual burden in SSA.

## Introduction

Female genital schistosomiasis (FGS) is a chronic and neglected gynaecological disease estimated to affect 30-56 million women globally, mostly in sub-Saharan Africa (SSA) (1,2). It is caused by the deposition of the eggs of *Schistosoma (S.) haematobium,* a waterborne trematode, in the female genital tract (1,2). The egg-deposition is a consequence of entrapment during egg migration from the pelvic venous plexus which induces an inflammatory response leading to the formation of granulomas, and epithelial damage presenting as characteristic cervicovaginal mucosal lesions (1,2). Untreated, FGS is associated with sexual dysfunction and reproductive tract morbidity, including infertility, dyspareunia and genital symptoms mimicking sexually transmitted infections (STIs) (2). Despite its high prevalence and clinical burden, FGS remains largely underdiagnosed and overlooked in schistosomiasis, and sexual and reproductive health programmes (1,2).

There are currently no guidelines for FGS screening, diagnosis, treatment, or case management, and awareness is low among communities and healthcare providers in many endemic settings (2,3,4). Conventional diagnosis involves collecting cervicovaginal images in clinics using a colposcope (2, 3,4). Images are then analysed by gynaecologists and classified as suggestive of *‘visual FGS* if characteristic cervicovaginal mucosal findings, including homogeneous sandy patches, grainy sandy patches, rubbery papules, or abnormal blood vessels are observed (5). Visual diagnosis of FGS lesions requires expensive equipment, advanced clinical infrastructures, and specialised training, which are often not available in *S. haematobium* endemic settings (2,6). In addition, concordance between expert reviewers is poor (Cohen’s kappa statistics=0.16), highlighting the low specificity and limitations of visual FGS diagnosis on cervicovaginal images (6). Molecular testing using polymerase chain reaction (PCR) to detect *S. haematobium* DNA in genital swabs (*‘molecular FGS’)* offers a potentially more scalable and specific diagnostic method (3,7). Genital samples can be collected in the clinic by a health worker or self-collected at home by an individual, potentially increasing screening coverage (3,7).

Cervical cancer incidence and mortality is highest among women in SSA (8–10). Zambia has the third highest incidence and mortality rates of cervical cancer in the world, with 65.5 new cases and 43.4 cancer deaths per 100,000 women per year (11). Persistent infection with one of the 12 high-risk (HR-) human papillomavirus (HPV) types (HPV types 16, 18, 31, 33, 35, 39, 45, 51, 52, 56, 58, and 59) is the primary cause of cervical cancer globally (12). HPV16 and 18 are responsible for 70% of all cervical cancer cases, while HPV31, 33, 45, 52, and 58 contribute to an additional 20% of cases (12,13). HR-HPV is primarily sexually transmitted, and risk factors include early sexual debut, greater number of lifetime sexual partners, high parity, hormonal contraceptive use, and co-infection with other STIs (14). Women living with HIV have a two-fold increased risk of acquiring HR-HPV (15), and a six-fold increased risk of cervical cancer, compared to women without HIV (16). The recovery of cell-mediated immunity following initiation of antiretroviral therapy (ART) does not necessarily remove this risk, although early initiation of ART before significant immune depletion reduces the risk of cervical cancer (17). The chronic genital inflammation caused by *S. haematobium* eggs may facilitate HR-HPV acquisition, persistence and progression to cervical precancer, yet this association requires further investigation (2,18).

The World Health Organization (WHO) has called for the elimination of cervical cancer by 2030, with targets including vaccinating 90% of girls by age 15; screening of 70% of women twice by age 35 and 45 with high-performance test; and treating 90% of women with cervical precancer and cancer (19). Zambia launched its national HPV vaccination programme in 2019 and is currently transitioning from visual inspection with acetic acid (VIA) to HPV DNA-based screening for both the general population of women and women living with HIV, due to its higher sensitivity and specificity to detect cervical precancer (20). HPV DNA testing can be performed on provider-or self-collected sample (20). Self-sampling for HPV screening has comparable sensitivity and specificity to clinician-collected samples when tested using PCR (21,22). Molecular testing for FGS could be integrated into the cervical cancer screening programmes and delivered at home for multi-pathogen community-based screening (7,8). Understanding the epidemiological overlap between FGS and HR-HPV infection is essential to inform integrated screening strategies (7).

Previous studies exploring the association of FGS and HPV in SSA showed mixed results (2,18). A study in Zimbabwe among 236 women of reproductive age (15-49 years) found evidence of an association between FGS, defined with the composite diagnosis of visual and molecular methods, and HR-HPV infection (age-adjusted OR=1.9, 95%CI 1.1-3.6, p-value=0.032) (23). Similarly, another cross-sectional study among 933 women in South Africa found strong evidence of an association between *visual FGS* by colposcopy and any HR-HPV infections (detected by GP5+/6+ HPV PCR test) (adjusted [a] OR: 1.71 95%

CI:1.14-2.56, p=0.010) (24). In contrast, a study of 302 women in Madagascar, found no evidence of an association between *visual FGS* and prevalence of any HPV infection (both high and low risk genotypes diagnosed using the E7-multiplex genotyping PCR assay) (OR=1.0, 95%CI 0.82-1.2) (25). A recent study in Tanzania, found no differences in rates of HR-HPV infection between women with and without *Schistosoma* infection, detected by circulating anodic antigen (CAA), an indirect measure of *Schistosoma* infection that does not indicate genital involvement (26).

To our knowledge no study to date has evaluated the association between FGS diagnosed using both *visual* and *molecular* diagnostic methods and individual HR-HPV genotypes. This study aims to determine the risk factors for HR-HPV and the association of HR-HPV prevalence and FGS diagnosed by *visual* and *molecular* diagnostic methods, in an ongoing longitudinal cohort in Zambia ( Zipime-Weka-Schista study).

## Methods

### Study design and participants

The *Zipime-Weka-Schista (Do-self-testing sister!)* study is a four-year longitudinal cohort study evaluating the performance and acceptability of multi-pathogen self-sampling for genital infections among women in Zambia, (27). This analysis uses baseline data collected between January 2022 and April 2023, across three sites (Livingstone, Sikoshwe, and Chanyanya). Briefly, girls and women aged from 15 to 50 years old, non-pregnant, sexually active, and who had been resident for at least a year in one of the study sites were eligible for enrolment. Participants were randomly selected through community-based random sampling and enrolled at home by trained Schista Community Workers (SCWs) (27). Further details are described in elsewhere (27).

### Samples collection

During the home visit, participants received study information, provided written informed consent, completed a questionnaire administerd by a SCW, and self-collected two cervicovaginal swabs and a urine specimen (27). One swab was stored in PrimeStore® MTM molecular transport media (donated by Longhorn Vaccines and Diagnostics LLC, Bethesda, US) for the detection of *Schistosoma* DNA (S1 Text) (27). The second swab was used for HR-HPV testing and was predominantly stored in PrimeStore® MTM molecular transport media (80% of baseline samples), with a smaller subset stored in ThinPrep Cytopreserve® solution (S1 Text). Urine was tested on the same day at the field laboratory for *S. haematobium* egg-patent infection by urinary egg microscopy (S2 Text). A 2mL urine sample was shipped to Leiden University Medical Centre (LUMC), The Netherlands for *Schistosoma* CAA detection using the up-converting reporter particle lateral flow CAA test (UCAA*hT*417 format, CAA-levels >0.6 pg/mL were considered positive) (S3 Text) (28). Participants were also offered HIV (OraQuick® HIV Self-Test) and *Trichomonas (T.) vaginalis* self-tests (OSOM® Trichomonas Rapid Test). For both self-tests, results were read by the SCW and immediately shared with the participant. HIV-positive self-test results were confirmed on-site using a point-of-care diagnostic test (Determine^TM^) (27). Participants who tested positive for HIV or *T. vaginalis* at home were referred to the clinic for treatment and linkage to care.

Following the home visit, enrolled participants who were not menstruating were invited for the clinic follow-up (27). After speculum insertion, a midwife collected two cervicovaginal swabs, a cervicovaginal lavage (CVL), and cervicovaginal images with a point-of-care colposcope (EVA MobileODT®, Tel Aviv, Israel), which are saved to an online portal for remote review. Similarly to the home visit, one cervicovaginal swab was stored in PrimeStore® MTM molecular transport (donated by Longhorn Vaccines and Diagnostics LLC, Bethesda, US) media and the second in Cytopreserve®. A CVL specimen was obtained by flushing 10mL of normal saline across the cervix and vaginal walls for one minute using a bulb syringe. The fluid was then collected from the posterior fornices with a pipette and transferred into a centrifuge tube containing PrimeStore® MTM. The CVL sample was stored at-20 °C on site (S1 Text).

### Visual FGS

Colposcopic cervicovaginal images were independently evaluated remotely by two experienced gynaecologists (BD and DK). In the case of disagreement between the two reviewers, a third reviewer (IH) was asked to independently analyse the images of the discordant case. Partcipant were defined as *‘visual FGS’* positive if at least two of the reviewers independently identified at least one of the four lesion types; homogeneous yellow sandy patches, grainy sandy patches, rubbery papules, or abnormal blood vessels, and as negative if none of these lesions were observed (5). Women with evidence of schistosomiasis, *‘molecular FGS’* and/or *‘visual FGS’* were treated with 40mg/kg praziquantel delivered as a single dose (30).

### Molecular FGS

Cervicovaginal swabs stored in Primestore® MTM were processed at Zambart central laboratory (Lusaka, Zambia) for the detection of *Schistosoma* DNA using Internal-trasncribed-spacer-2 (ITS-2) by real-time PCR (ITS qPCR) (S4 Text). Total nucleic acid (TNA) extraction (PrimeXtract™ Longhorn Vaccines & Diagnostics, USA) was performed as per manufacturer’s instructions, using a simplified silica-based spin column extraction process. qPCR was performed as previously reported and results were reported in cycle threshold values (Ct-values) and considered positive if any C_t_-value was observed and negative if no C_t_-value was observed (3,29). Women were classified as *‘molecular FGS’* positive if either self-or provider-collected cervicovaginal swab tested positive for *Schistosoma* DNA via qPCR.

### HR-HPV detection

Self-collected cervicovaginal swabs stored in either PrimeStore® Molecular Transport Medium (MTM) or in ThinPrep Cytopreserve® were tested for HR-HPV detection using a multiplex real-time PCR assay (GeneXpert HPV®, Cepheid, Sunnyvale, CA, USA) which simultaneously detects 14 HR-HPV types (HPV 16, 18, 45, 31, 33, 35, 52, 58, 51, 59, 39, 56, 66, 68), grouped into three channels: HPV type 16; HPV types 18 or 45 (or both); and any other HR-HPV types. DNA extraction for samples stored in PrimeStore® Molecular Transport Medium (MTM) was performed using a modified PrimeXtract™ spin column protocol (S5 Text). For GeneXpert HPVanalysis, 50 µL of the extracted DNA was diluted in 1mL of normal saline and loaded into the GeneXpert cartridge, following the standard assay protocol. A sample adequacy control (SAC) and a Probe Check Control (PCC) was also provided in the cartride (S5 Text). If a test result on the self-collected swab was invalid, HR-HPV testing was done using the healthcare provider-collected swab (S6 Text).

### HR-HPV definitions

Participants were classified as HR-HPV positive if positive for at least one of the 14 HR-HPV types (HPV 16, 18, 45, 31, 33, 35,52,58,51,39, 56, 66, 68); HPV16/18/45 positive if positive for the HPV type 16, and/or the HPV type 18 and/or 45 channels; and HPV16 positive if only the HPV type 16 channel was positive (S1 Figure). Samples were classified as invalid if both the home-based self-collected swab and clinic-based provider-collected swab returned invalid results (S6 Text). This classification allowed to distinguish HPV16 and HPV18 and HPV45 from the other HR-HPV genotypes as they are the most oncogenic genotypes (12,13). HR-HPV positive women were referred to a gynaecologist for colposcopy and a two-quadrant cervical biopsy according to age and HIV status (27). All women with cervical precancer were referred for management as per national guidelines (20).

### Statistical analyses

Data were entered on hand-held electronic devices using Open Data Kit (ODK) and analysed using Stata 17.0 (StataCorp, LP, College Station, US). Primary outcomes were detection of any HR-HPV, HPV type 16 or 18 and/or 45 (HPV16/18/45) and HPV type 16 only. The HPV16/18/45 outcome was defined by combining outputs from the HPV type 16, and the HPV type 18 and/or 45 channels. The denominator throughout this analysis was all women with valid HPV DNA test results. Descriptive statistics were used to summarise participants’ characteristics, stratified by HR-HPV status. Univariable logistic regression analyses were used to estimate crude odds ratios (cOR) and 95% confidence intervals (CI) for the associations between HR-HPV and potential risk factors. Potential risk factors were informed by the published literature. Multivariable logistic regression models were used to estimate adjusted odds ratios (aOR) and 95% CI for the associations between HR-HPV and factors associated with HR-HPV in univariable analysis. (p-value<0.05). Covariates with missing data or those applicable only to a subgroup were excluded. Multicollinearity was assessed between variables in the final models.

### Ethical considerations

The study was approved by the University of Zambia Biomedical Research Ethics Committee (UNZABREC) (reference: 1858-2021), the National Health Research Authority (NHRA) (Reference: 00012/24/09/2021), and the London School of Hygiene and Tropical Medicine (LSHTM) (reference: 25258). Ministry of Health and local superintendents approved the study in September 2021.

## Results

Figure 1 shows the study flow. At baseline, 2,532 women (median age: 28 years, interquartile range [IQR] 22-36) were enrolled at home, and 67% (1,694/2,532) completed the clinic follow-up after a median of two days (IQR: 1-11 days). Cervicovaginal swabs and urine samples were collected from all women at enrolment. All urine samples were tested for *S. haematobium* ova detection by microscopy, and 99.4% (2,517/2,532) were tested for presence of CAA. *Visual FGS* status was available for 99.8% (1,691/1,694) of participants who completed the clinic follow-up. HR-HPV DNA testing was performed on 98.3% (2,488/2,532) of participants, and 96.5% (2,401/2,488) gave a valid HR-HPV result. *Molecular FGS* diagnosis was available for all study participants (Figure 1). Data for both HR-HPV and *molecular FGS* status were available for 94.8% (2,401/2,532) of participants, and for HR-HPV and *visual FGS* in 96.7% (1,639/1,694) of clinic attendees.

**Figure 1:**
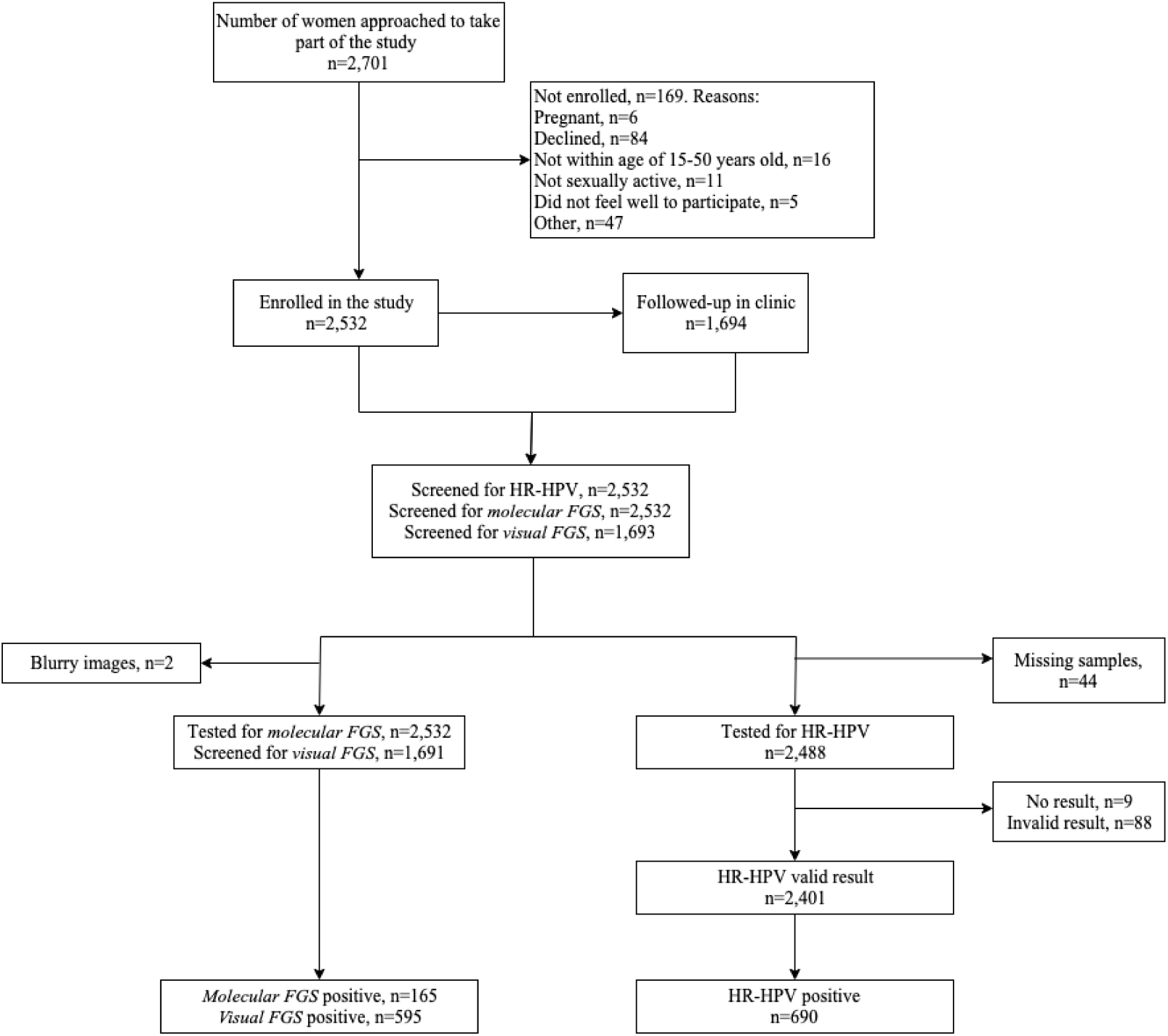
*Zipime-Weka-Schista* study flow diagram for participant recruitment at baseline and screening of *molecular FGS, visual FGS* and HR-HPV.

### Participant characteristics

Of 2,532 women, 165 (6.5%) tested positive for *molecular FGS* (median Ct-value= 35.1, IQR:7.4-31.6) (Table 1). *Visual FGS* was identified in 35.2% (595/1,691) of participants, (Table 1). Egg-patent *S. haematobium* infection was detected in 5.2% (132/2,532) of participants (mean intensity: 4.8 eggs/10mL of urine; Standard deviation [SD]: 5.5). *Schistosoma* CAA positivity was observerd in 15.4% (388/2,517) (median CAA concentration=4.6 pg/mL; IQR 1.1-31.0) (Table 1). HIV prevalence was 17.5% (443/2,532), with 91.6% (406/443) of self-reporting antiretroviral therapy (ART) use (Table 1). The prevalence of *T. vaginalis* by rapid diagnostic test (RDT) was 10.3% (246/2,389) (Table 1).

**Table 1:**
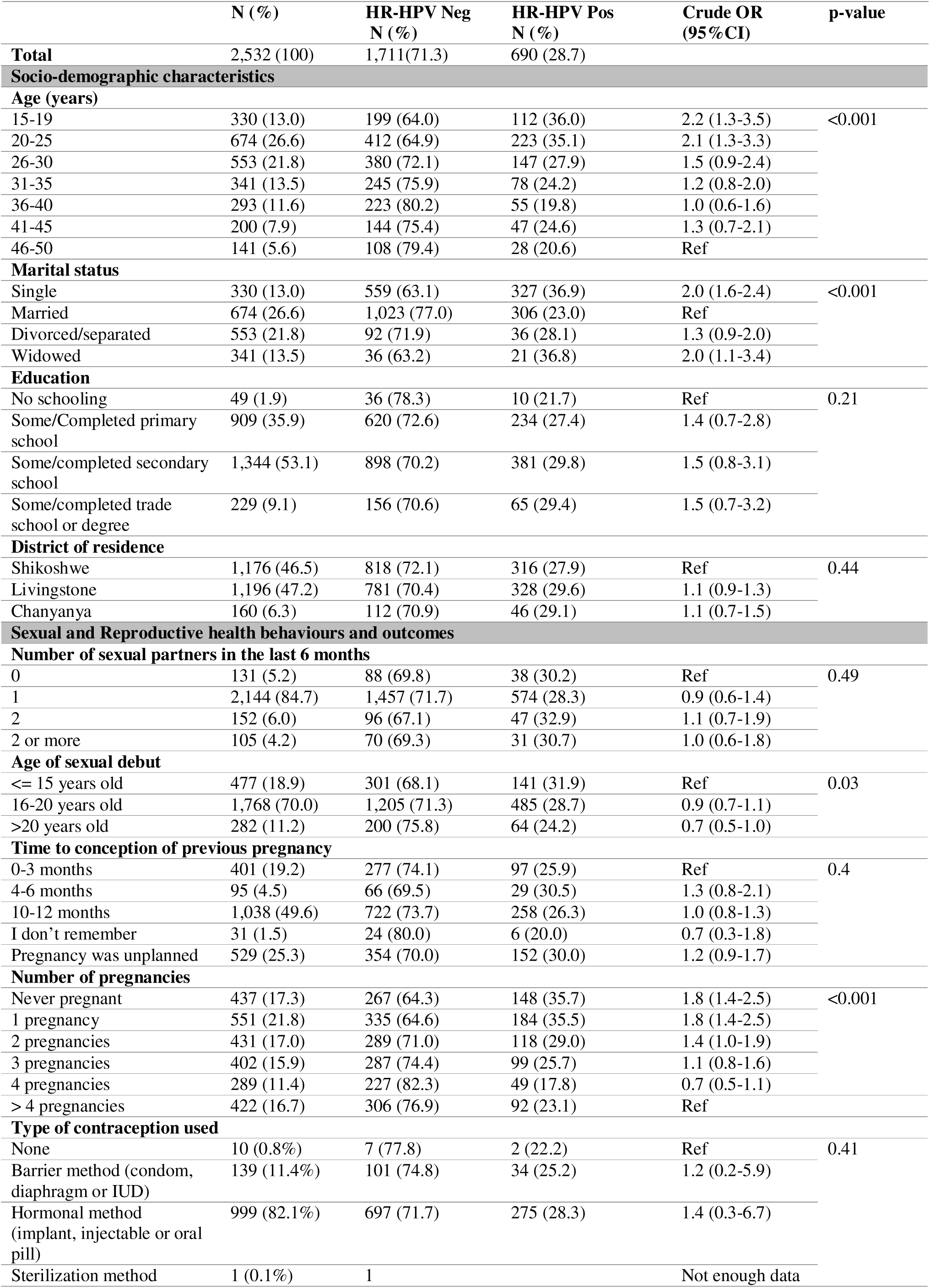
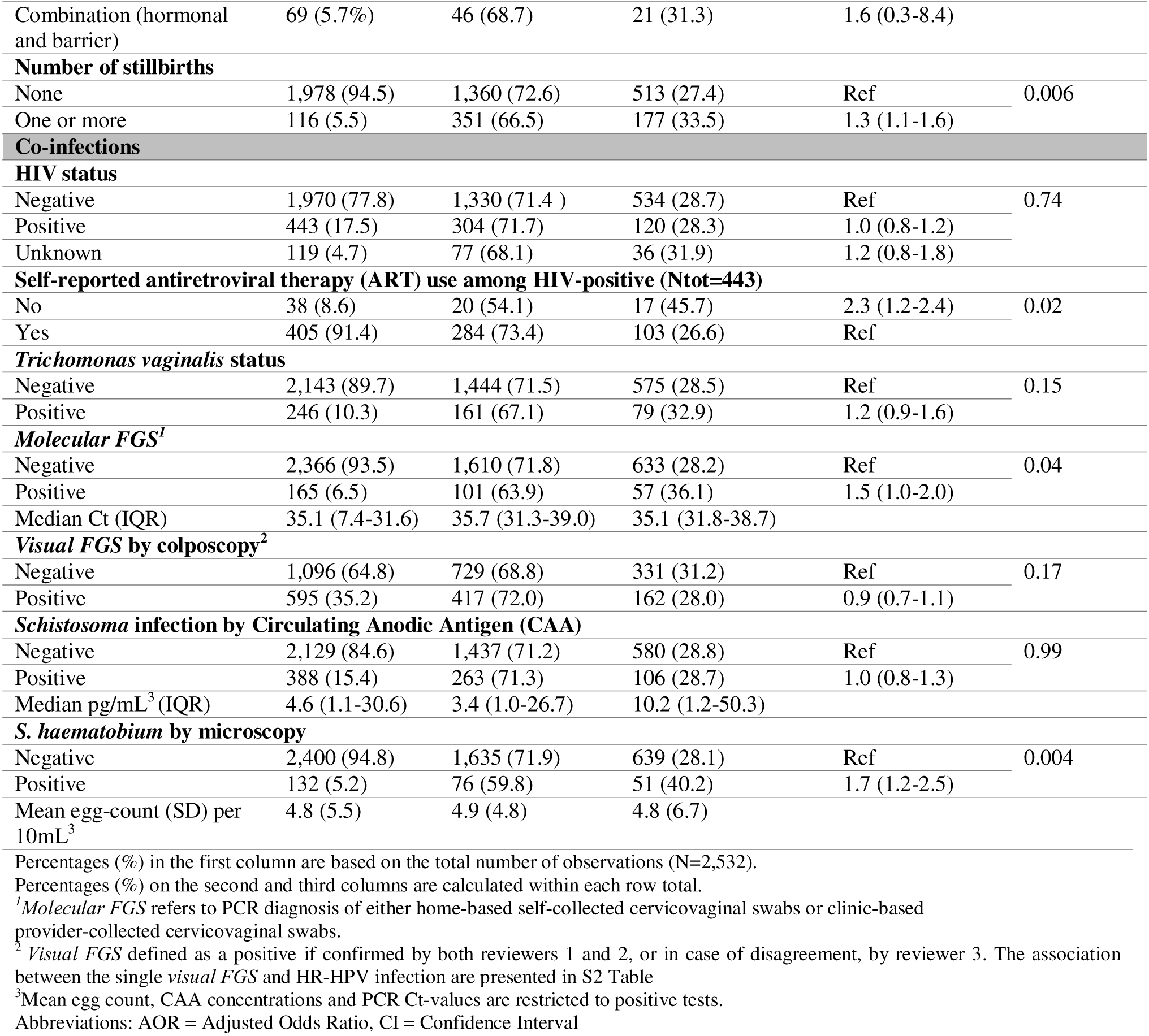
Prevalence of any high-risk HPV (HR-HPV) stratified by socio-demographic characteristics and co-infections, with univariable logistic regression estimates across the study population (N = 2,401).

### Distribution of infection by age

The distribution of *molecular FGS, visual FGS* and HR-HPV infection by age is shown in Figure 2 (S1 Table). The prevalence of *molecular FGS* was highest among adolescents aged 15-19 years (14.6%) and declined with age (χ² < 0.00) (Figure 2, S1 Table). In contrast, prevalence of *visual FGS* increased with age, peaking among women aged 46-50 years (53.4%) χ² < 0.001) (Figure 2, S1 Table). HR-HPV prevalence declined significantly with age (p-value<0.001). Compared to women aged 46-50 years, girls and women aged 15-25 years old had higher risk of HR-HPV (15-19 years vs. 46-50 years; 36.0% vs. 20.6%; cOR =2.2, 95%CI 1.3-3.5; 20-25 years vs. 46-50 years: 35.1% cOR=2.1, 95% CI 1.3-3.3) (Figure 2 and Table 1).

**Figure 2:**
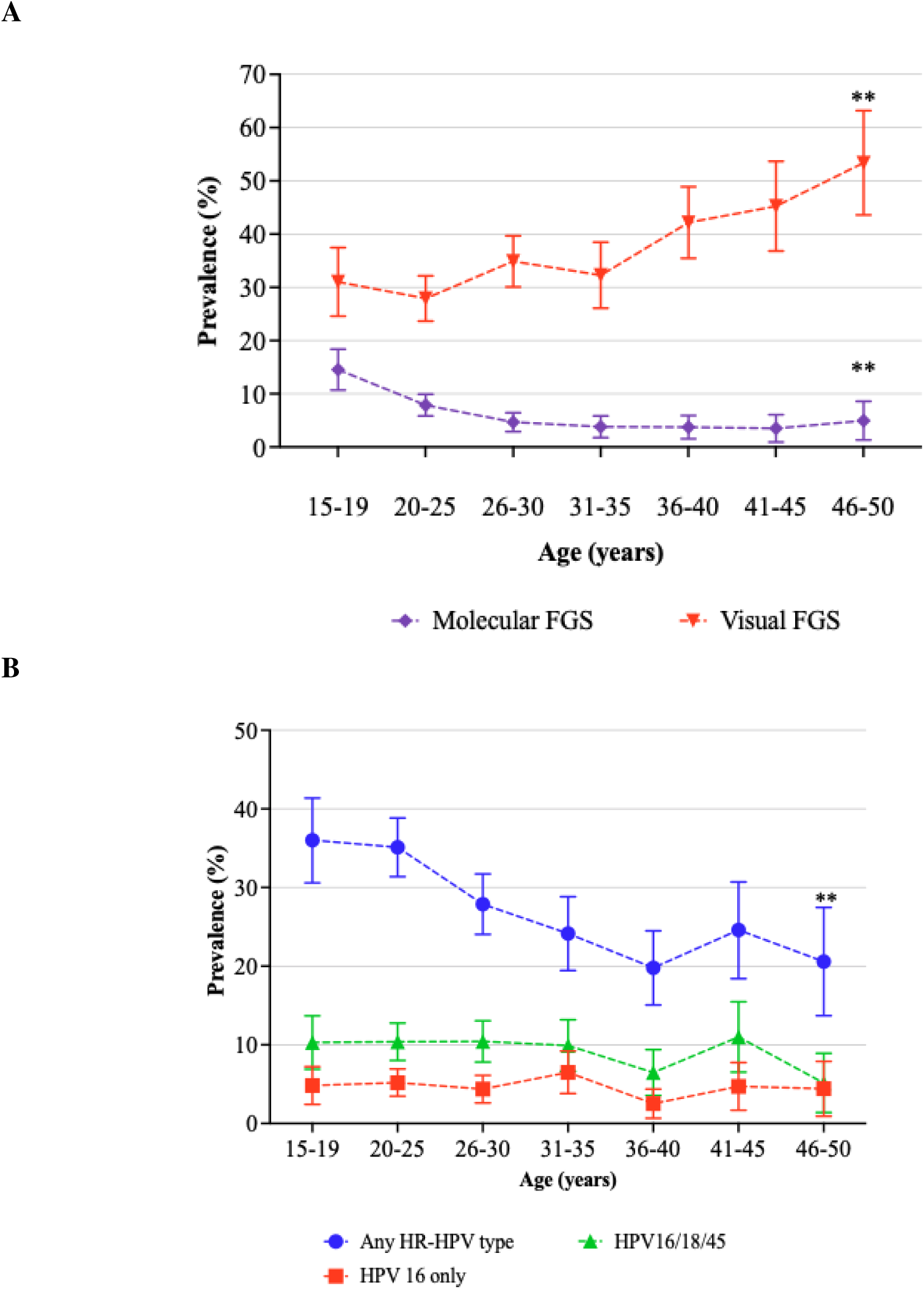
Distribution of positive diagnostic test results by age categories. **A**: Distribution of molecular FGS and visual FGS by age groups. **B**: Prevalence of any HR-HPV infection, HPV16/18/45 and HPV16 only by age categories. The corresponding data is presented in Table 1 and S1 Table.

### Prevalence and risk factor analysis for prevalence of *any* HR-HPV

Prevalence of any HR-HPV genotypes was 28.7% (690/2,401) (Table 1). HR-HPV prevalence declined significantly with age (p-value<0.001). Any HR-HPV prevalence was higher among women with *molecular FGS* compared to those without (36.1% vs. 28.2%; cOR= 1.5, 95%CI 1.0-2.0) (Table 1). No statistically significant difference was observed by *visual FGS* status (*visual FGS* positive vs. negative 28.0% vs. 31.2%, cOR=0.9, 95%CI 0.7-1.1), either overall or by lesion type (Table 1 and S2 Table). Women with urinary *S. haematobium* by microscopy had a higher prevalence of any HR-HPV compared to those without (40.2% vs. 28.1, cOR=1.7, 95%CI 1.2-2.5). However, this association was not observed with *Schistosoma* infection by CAA (cOR=1.0, 95%CI 0.8-1.3) (Table 1). No significant differences in HR-HPV prevalence were observed by HIV status (cOR=1.0, 95%CI 0.8-1.2) or *T. vaginalis* status (cOR= 1.2, 95%CI 0.9-1.6) (Table 1). After adjusting for age, marital status, and number of pregnancies *molecular FGS* was not significantly associated with any HR-HPV prevalence (aOR=1.3, 95% CI=0.9-1.9) (Figure 3). Women aged 20-25 years had higher odds of HR-HPV infection compared to women aged >= 45 years (aOR=1.65, 95%CI 1.1-2.5). Married women had lower odds of HR-HPV infection compared to single women (aOR=0.6, 95%CI 0.5-0.8) (Table 1, Figure 3. Due to multicollinearity between *‘molecular FGS’* and urinary *S. haematobium* infection (r=0.56), a separate model evaluated the association between urinary *S. haematobium* infection by microscopy and infection with any HR-HPV (aOR=1.7, 95%CI 1.1–2.4) (S3 Table)

**Figure 3:**
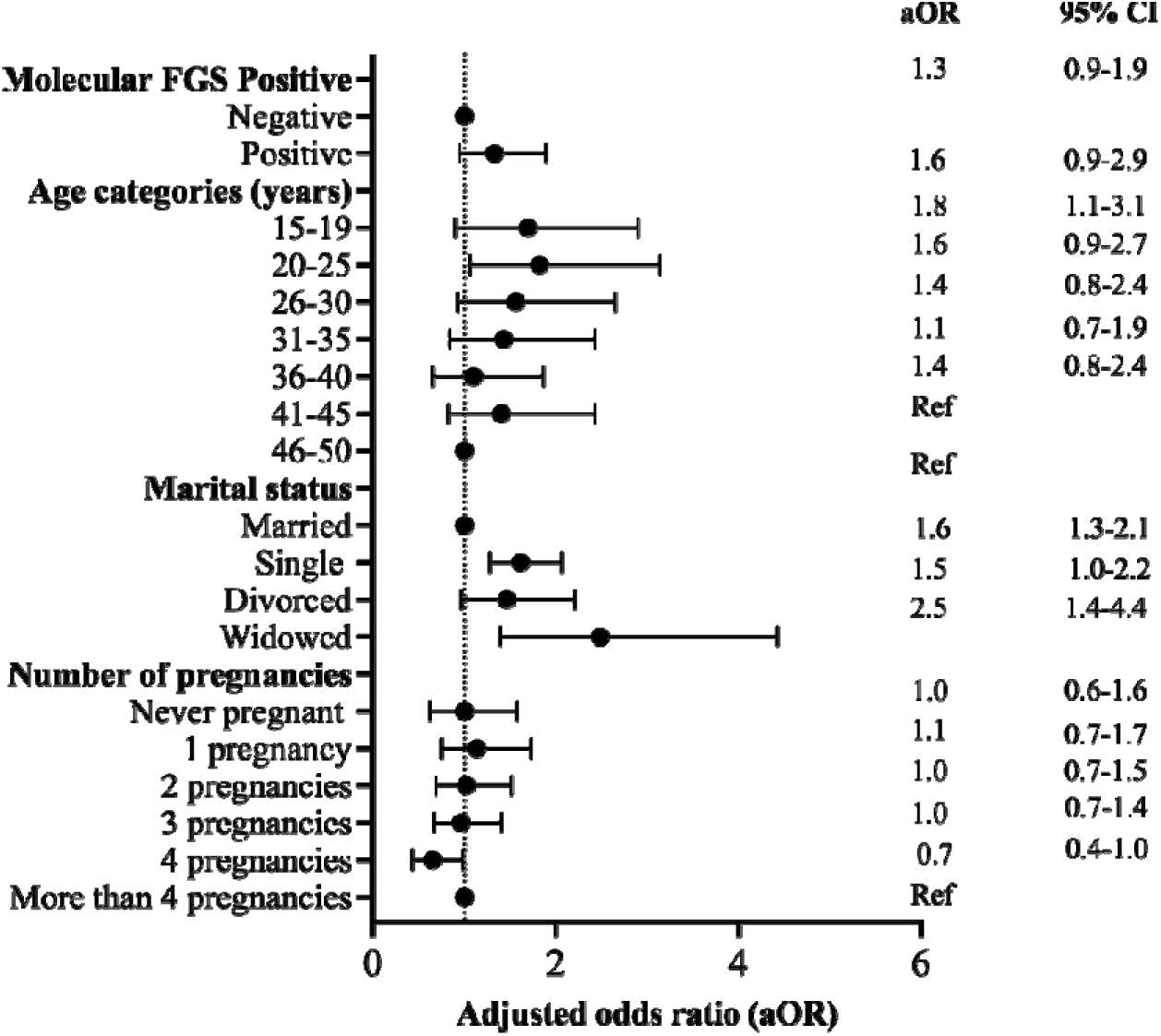
Multivariable logistic regression model parameter estimates with prevalence of any HR-HPV as the dependent variable across the study population (N=2,401). The regression model was adjusted for molecular FGS status, age, marital status and number of pregnancies. Abbreviations: aOR= Adjusted odds ratios.

### Prevalence and risk factor analysis for infection with high oncogenic types; HPV16, 18 and 45 HPV 16/18/45

Prevalence of infection with any of the HPV16/18/45 was 9.6% (231/2,401) (Table 2). In univariable analysis, higher prevalence was observed among HIV-positive versus HIV-negative women (12.5% [53/424] vs. 8.9% [166/1,864], cOR=1.5, 95%CI 1.1-2.0), and among women with *T. vaginalis* compared to those without (14.6% [35/240] vs. 8.8% [177/2,019], cOR=1.8, 95%CI 1.2-2.6) (Table 2). HPV16/18/45 prevalence was higher among women with *molecular FGS* compared to those without (15.8% [25/158] vs. 9.2% [206/2,243], cOR=1.9, 95%CI 1.2-2.9), but no association was observed with *visual FGS* (cOR=0.8, 95%CI 0.6-1.2) (Table 2, S4 Table). In adjusted analysis, *molecular FGS* (aOR=1.7, 95%CI 1.0-2.8), HIV status (aOR=1.7, 95%CI 1.2-2.4), and *T. vaginalis* status (aOR=1.5, 95%CI 1.0-2.2) were associated with increased odds of HPV16/18/45 infection (Figure 4).In addition, single women had higher odds of HPV16/18/45 infection compared to married women, (aOR=1.9, 95%1.4-2.9), and participants residing in Chanyanya were more likely to be infected than those from Shikoswe (aOR=3.6, 95%CI 1.4-9.3) (Figure 4).

**Figure 4:**
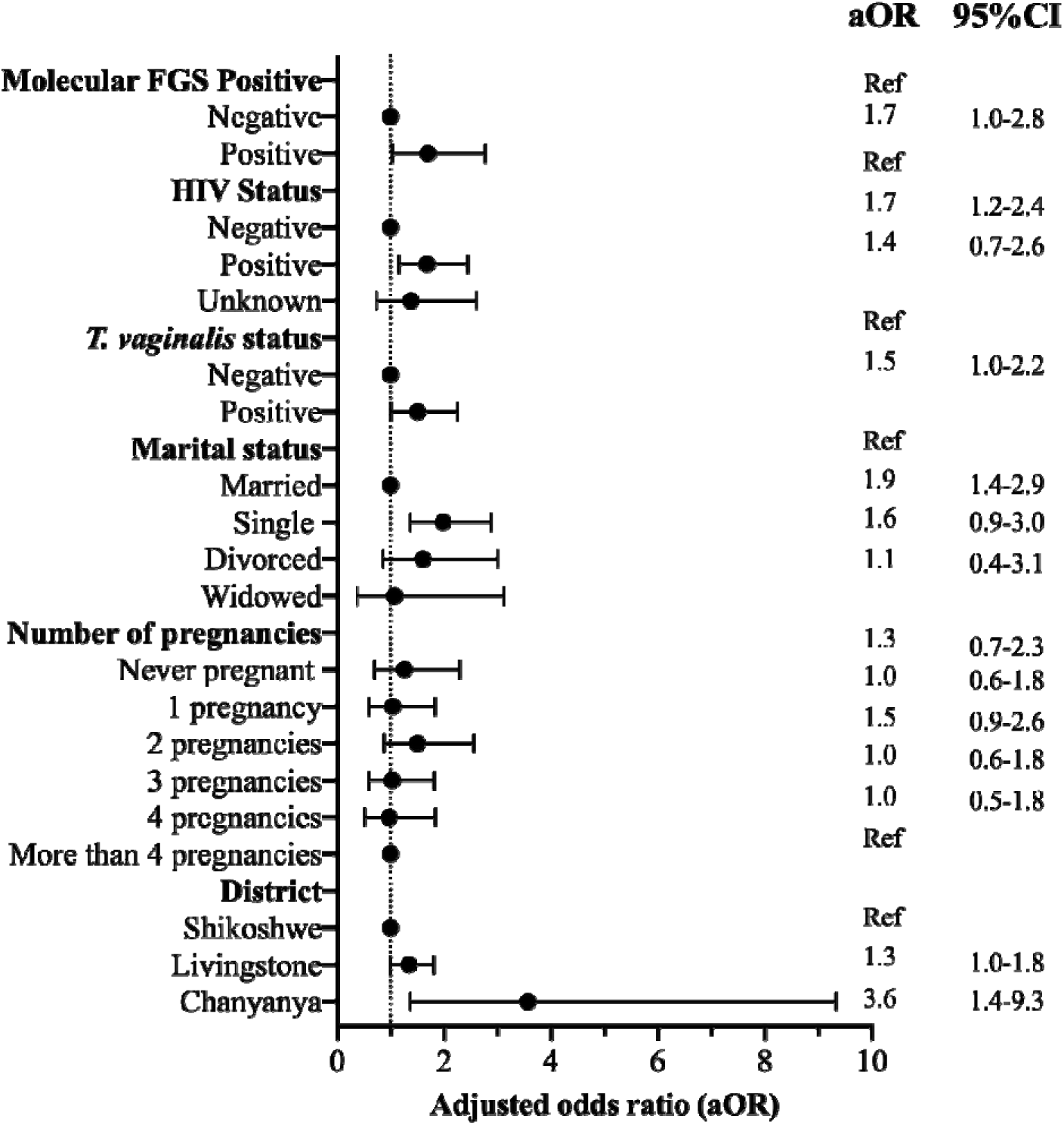
Multivariable logistic regression model parameter estimates for prevalence of HPV16/18/45 as the dependent variable in the study population (N=2,401). The model was adjusted for *molecular FGS* status, HIV status, *T. vaginalis* status, district of residence, marital status and number of pregnancies. Abbreviations: aOR=Adjusted Odds Ratio, CI=Confidence Intervals.

**Table 2:** Prevalence of HPV16/18/45 stratified by socio-demographic characteristics and co-infections, with univariable logistic regression estimates across the study population (N = 2,401).

|  | HPV16/18/45 Neg N (%) | HPV16/18/45 Pos N (%) | Crude OR (95%CI) | p-value |
| --- | --- | --- | --- | --- |
| Total | 2,170 (90.4) | 231 (9.6) |  |  |
| Socio-demographic characteristics |  |  |  |  |
| Age (years) |  |  |  |  |
| 15-19 | 279 (89.7) | 32 (10.3) | 2.1 (0.91-4.9) | 0.09 |
| 20-25 | 569 (89.6) | 66 (10.4) | 2.1 (1.0-4.8) |  |
| 26-30 | 472 (89.6) | 55 (10.4) | 2.1 (1.0-4.8) |  |
| 31-35 | 291 (90.1) | 32 (9.9) | 2.0 (0.9-4.7) |  |
| 36-40 | 260 (93.5) | 18 (6.5) | 1.3 (0.5-3.1) |  |
| 41-45 | 170 (89.0) | 21 (11.0) | 2.3 (0.9-5.1) |  |
| 46-50 | 129 (94.9) | 7 (5.2) | Ref |  |
| Marital status |  |  |  |  |
| Single | 770 (86.9) | 116 (13.1) | 1.9 (1.5-2.6) | 0.001 |
| Married | 1,233 (92.8) | 96 (7.2) | Ref |  |
| Divorced/<br>separated | 113 (88.3) | 15 (11.7) | 1.7 (1.0-3.0) |  |
| Widowed | 53 (93.0) | 4 (7.0) | 1.0 (0.3-2.7) |  |
| Education |  |  |  |  |
| No schooling | 44 (95.7) | 2 (4.4) | Ref | 0.87 |
| Some/Completed primary school | 769 (90.1) | 85 (9.9) | 2.4 (0.6-10.2) |  |
| Some/completed secondary school | 1,157 (90.5) | 122 (9.5) | 2.3 (0.6-9.7) |  |
| Some/completed trade school or degree | 199 (90.1) | 22 (10.0) | 2.4 (0.6-10.7) |  |
| District of residence |  |  |  |  |
| Shikoshwe | 1,047 (92.3) | 87 (7.7) | Ref | <0.001 |
| Livingstone | 990 (89.3) | 119 (10.7) | 1.4 (1.1-1.9) |  |
| Chanyanya | 133 (84.2) | 25 (15.8) | 2.3 (1.4-3.7) |  |
| Sexual and Reproductive health behaviours and outcomes |  |  |  |  |
| Number of sexual partners in the last 6 months |  |  |  |  |
| 0 | 115 (91.3%) | 11 (8.7%) | Ref | 0.60 |
| 1 | 1,836 (90.4%) | 195 (9.6%) | 1.1 (0.6-2.1) |  |
| 2 | 129 (90.2%) | 14 (9.8%) | 1.1 (0.5-2.6) |  |
| More than 2 | 90 (89.1%) | 11 (10.9%) | 1.3 (0.5-3.0) |  |
| Age of sexual debut |  |  |  |  |
| <= 15 years old | 389 (88.0%) | 53 (12.0%) | Ref | 0.16 |
| 16-20 years old | 1,537 (91.0%) | 153 (9.1%) | 0.7 (0.5-1.0) |  |
| >20 years old | 239 (90.5%) | 25 (9.5%) | 0.8 (0.5-1.3) |  |
| Time to conception of previous pregnancy |  |  |  |  |
| 0-3 months | 350 (93.6) | 24 (6.4) | Ref | 0.26 |
| 4-6 months | 85 (89.5) | 10 (10.5) | 1.7 (0.8-3.7) |  |
| 10-12 months | 891 (90.9) | 89 (9.1) | 1.5 (0.9-2.3) |  |
| I don't remember | 29 (93.3) | 2 (6.7) | 1.0 (0.2-4.6) |  |
| Pregnancy was unplanned | 452 (89.3) | 54 (10.7) | 1.7 (1.1-2.9) |  |

| Type of contraception used |  |  |  |  |
| --- | --- | --- | --- | --- |
| None | 78 (86.7%) | 12 (13.3%) | Ref | 0.50 |
| Barrier method (condom, diaphragm or IUD) | 22 (91.7%) | 2 (8.3%) | 0.6 (0.1-2.8) |  |
| Hormonal method (implant, injectable or oral pill) | 254 (90.1%) | 28 (9.9%) | 0.7 (0.3-1.5) |  |
| Sterilization method+ | 1 (100%) | 0 | Not enough data |  |
| Combination (hormonal and barrier) | 681 (90.3%) | 73 (9.7%) | 0.7 (0.4-1.3) |  |
| Number of pregnancies |  |  |  |  |
| Never pregnant | 363 (87.5) | 52 (12.5) | 1.9 (1.1-3.4) | 0.01 |
| 1 pregnancy | 469 (90.4) | 50 (9.6) | 1.4 (0.8-2.5) |  |
| 2 pregnancies | 361 (88.7) | 46 (11.3) | 1.7 (1.0-3.0) |  |
| 3 pregnancies | 355 (92.0) | 31 (8.0) | 1.2 (0.7-2.1) |  |
| 4 pregnancies | 257 (93.1) | 19 (6.9) | 1.2 (0.7-2.2) |  |
| > 4 pregnancies | 365 (91.7) | 33 (8.3) | Ref |  |
| Number of stillbirths |  |  |  |  |
| None | 1,708 (91.1) | 166 (8.9) | Ref | 0.02 |
| One or more | 463 (87.7) | 65 (12.3) | 1.4 (1.1-2.0) |  |
| Co-infections |  |  |  |  |
| Molecular FGS<sup>1</sup> |  |  |  |  |
| Negative | 2,037 (90.8) | 206 (9.2) | Ref | 0.007 |
| Positive | 133 (84.2) | 25 (15.8) | 1.9 (1.2-2.9) |  |
| Visual FGS by colposcopy<sup>2</sup> |  |  |  |  |
| Negative | 947 (89.3) | 113 (10.7) | Ref | 0.28 |
| Positive | 527 (91.0) | 52 (9.0) | 0.8 (0.6-1.2) |  |
| HIV status |  |  |  |  |
| Negative | 1,698 (91.1) | 166 (8.9) | Ref | 0.07 |
| Positive | 371 (87.5) | 53 (12.5) | 1.5 (1.1-2.0) |  |
| Unknown | 101 (89.4) | 12 (10.6) | 1.2 (0.7-2.3) |  |
| Self-reported antiretroviral therapy (ART) use among HIV-positive |  |  |  |  |
| No | 30 (81.1) | 7 (18.9) | 1.8 (0.7-4.2) | 0.22 |
| Yes | 341 (88.1) | 46 (11.9) | Ref |  |
| Trichomonas (T.) vaginalis status (Ntot=1,556) |  |  |  |  |
| Negative | 1,842 (91.2) | 177 (8.8) | Ref | 0.004 |
| Positive | 205 (85.4) | 35 (14.6) | 1.8 (1.2-2.6) |  |
| S. haematobium by Circulating Anodic Antigen (CAA) (Ntot=1,685) |  |  |  |  |
| Negative | 1,829 (90.7) | 188 (9.3) | Ref | 0.22 |
| Positive | 327 (88.6) | 42 (11.4) | 1.2 (0.9-1.8) |  |
| S. haematobium by urine microscopy (Ntot=1,691) |  |  |  |  |
| Negative | 2,063 (90.7) | 211 (9.3) | Ref | 0.02 |
| Positive | 107 (84.3) | 20 (15.8) | 1.8 (1.1-3.0) |  |

Percentages (%) on the second and third columns are calculated within each row total.
<sup>1</sup>*Molecular FGS* refers to PCR diagnosis of either home-based self-collected cervicovaginal swabs or clinic-based provider-collected cervicovaginal swabs.
<sup>2</sup>*Visual FGS* defined as a positive if confirmed by both reviewers 1 and 2, or in case of disagreement, by reviewer 3. The association between the single *visual FGS* and infection with HPV16/18/45 are presented in S4 Table

### HPV16 only

HPV16 prevalence was 4.8% (114/2,401) (Table 3). HPV16 prevalence was higher among single women compared to married women (6.6% [58/886] vs. 3.5% [46/1,329], cOR=2.0, 95%CI 1.3-2.9) (Table 3). Women with *molecular FGS* had higher HPV16 prevalence compared to those without (8.2% [13/158] vs. 4.5% [101/2,243]; cOR=1.9, 95%CI 1.0-3.5) (Table 3). No significant association was observed with *visual FGS* status (cOR=0.8, 95%CI 0.5-1.2), urinary *S. haematobium* status by microscopy (cOR=1.4, 95%CI 0.7-2.9), *Schistosoma* status by CAA (cOR=1.2, 95%CI 0.8-2.0), HIV status (cOR=1.4, 95%CI 0.9-2.2), or *T. vaginalis* status (cOR=1.4, 95%CI 0.8-2.5) (Table 3, S5 Table). After adjusting for marital status and parity, women with *molecular FGS* had 1.8 higher odds of being HPV16 positive compared to those without (aOR=1.8, 95%CI 1.0-3.4) (Figure 5).

**Figure 5:**
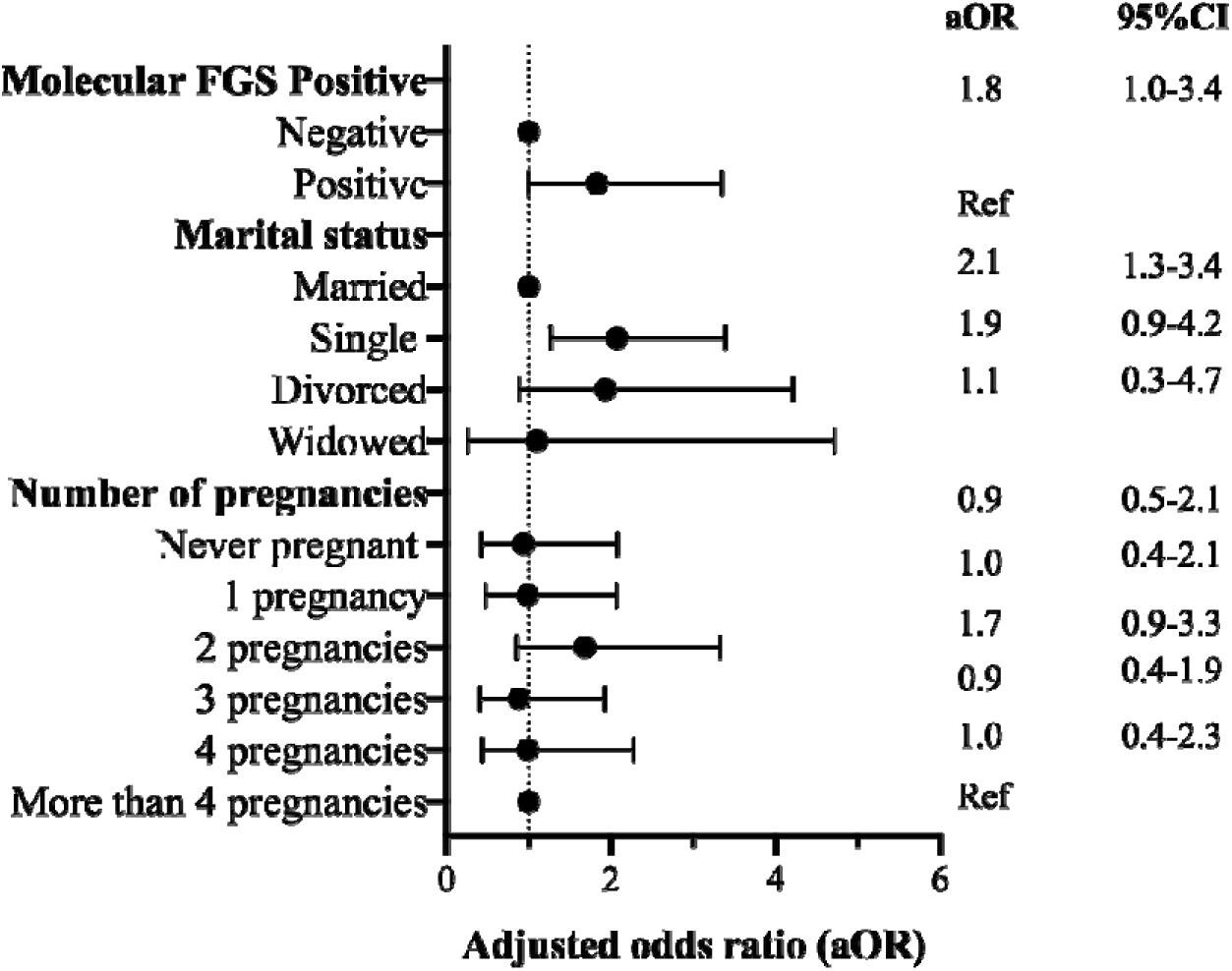
Multivariable logistic regression model parameter estimates for prevalence of HPV16 as the dependent variable in the study population (N=2,401). The logistic regression model was adjusted for *molecular FGS* status, marital status and number of pregnancies. Abbreviations: aOR= Adjusted odds ratio, CI= Confidence Intervals

**Table 3:** Prevalence of HPV16 only stratified by socio-demographic characteristics and co-infections, with univariable logistic regression estimates across the study population (N = 2,401).

|  | HPV16 Neg<br>N (%) | HPV16 Pos<br>N (%) | Crude OR (95%CI) | p-value |
| --- | --- | --- | --- | --- |
| Total | 2,287 (95.3) | 114 (4.8) |  |  |
| Socio-demographic characteristics |  |  |  |  |
| Age (years) |  |  |  |  |
| 15-19 | 296 (95.2) | 15 (4.8) | 1.1 (0.4-2.9) | 0.50 |
| 20-25 | 602 (94.8) | 33 (5.2) | 1.2 (0.5-2.9) |  |
| 26-30 | 504 (95.6) | 23 (4.4) | 1.0 (0.4-2.5) |  |
| 31-35 | 302 (93.5) | 21 (6.5) | 1.5 (0.6-3.8) |  |
| 36-40 | 271 (97.5) | 7 (2.5) | 0.6 (0.2-1.7) |  |
| 41-45 | 182 (95.3) | 9 (4.7) | 1.1 (0.4-3.1) |  |
| 46-50 | 130 (95.6) | 6 (4.4) | Ref |  |
| Marital status |  |  |  |  |
| Single | 828 (93.5) | 58 (6.6) | 2.0 (1.3-2.9) | 0.03 |
| Married | 1,283 (96.5) | 46 (3.5) | Ref |  |
| Divorced/<br>separated | 120 (93.8) | 8 (6.3) | 1.9 (0.9-4.0) |  |
| Widowed | 55 (96.5) | 2 (3.5) | 1.0 (0.2-4.3) |  |
| Education |  |  |  |  |
| No schooling | 45 (97.8) | 1 (2.2) | Ref | 0.38 |
| Some/Completed primary school | 815 (95.4) | 39 (4.6) | 2.2 (0.3-16.0) |  |
| Some/completed secondary school | 1,218 (95.2) | 61 (4.8) | 2.3 (0.3-16.6) |  |
| Some/completed trade school or<br>degree | 208 (94.1) | 13 (5.9) | 2.8 (0.4-22.1) |  |
| District of residence |  |  |  |  |
| Shikoshwe | 1,091 (96.2) | 43 (3.8) | Ref | 0.055 |
| Livingstone | 1,047 (94.4) | 62 (5.6) | 1.5 (1.0-2.2) |  |
| Chanyanya | 149 (94.3) | 9 (5.7) | 1.5 (0.7-3.2) |  |
| Sexual and Reproductive Health Behaviours and Outcomes |  |  |  |  |
| Number of sexual partners in the last 6 months |  |  |  |  |
| 0 | 118 (94.4%) | 7 (5.6%) | Ref | 0.94 |
| 1 | 1,937 (95.4%) | 94 (4.6%) | 0.8 (0.4-1.8) |  |
| 2 | 134 (93.7%) | 9 (6.3%) | 1.1 (0.4-3.2) |  |
| More than 2 | 97 (96.0%) | 4 (4.0%) | 0.7 (0.2-2.5) |  |
| Age of sexual debut |  |  |  |  |
| <= 15 years old | 419 (94.8%) | 23 (5.2%) | Ref | 0.18 |
| 16-20 years old | 1,606 (95.0%) | 84 (5.0%) | 1.0 (0.6-1.5) |  |
| >20 years-old | 257 (97.4%) | 7 (2.7%) | 0.5 (0.2-1.2) |  |
| Time to conception |  |  |  |  |
| 0-3 months | 363 (97.1) | 11 (2.9) | Ref | 0.28 |
| 4-6 months | 90 (94.7) | 5 (5.3) | 1.8 (0.6-5.4) |  |
| 10-12 months | 938 (95.7) | 42 (4.3) | 1.5 (0.8-2.9) |  |
| I don't remember | 28 (93.3) | 2 (6.7) | 2.4 (0.5-11.2) |  |
| Pregnancy was unplanned | 476 (94.1) | 30 (5.9) | 2.1 (1.0-4.2) |  |
| Type of contraception used |  |  |  |  |
| None | 84 (93.3%) | 6 (6.7%) | Ref | 0.72 |
| Barrier method (condom, diaphragm or IUD) | 23 (95.8%) | 1 (4.2%) | 0.6 (0.07-5.3) |  |
| Hormonal method (implant, injectable or oral pill) | 268 (95.4%) | 13 (4.6%) | 0.7 (0.2-1.8) |  |
| Sterilization method+ | 1 (100%) |  | No data |  |
| Combination (hormonal and barrier) | 714 (94.7%) | 40 (5.3%) | 0.8 (0.32-1.9) |  |
| **Number of pregnancies** |  |  |  |  |
| Never pregnant | 391 (94.2) | 24 (5.8) | 1.7 (0.9-3.3) | 0.04 |
| 1 pregnancy | 493 (95.0) | 26 (5.0) | 1.4 (0.7-2.8) |  |
| 2 pregnancies | 380 (93.4) | 27 (6.6) | 1.9 (1.0-3.8) |  |
| 3 pregnancies | 373 (96.6) | 13 (3.4) | 1.0 (0.4-2.1) |  |
| 4 pregnancies | 266 (96.4) | 10 (3.6) | 1.0 (0.5-2.4) |  |
| > 4 pregnancies | 384 (96.5) | 14 (3.5) | Ref |  |
| **Number of stillbirths** |  |  |  |  |
| None | 1,789 (95.5) | 84 (4.5) | Ref | 0.25 |
| One or more | 498 (94.3) | 30 (5.7) | 1.3 (0.8-2.0) |  |
| **Co-infections** |  |  |  |  |
| ***Molecular FGS<sup>1</sup>*** |  |  |  |  |
| Negative | 2,142 (95.5) | 101 (4.5) | Ref | 0.04 |
| Positive | 145 (91.8) | 13 (8.2) | 1.9 (1.0-3.5) |  |
| ***Visual FGS by point-of-care colposcopy<sup>2</sup>*** |  |  |  |  |
| Negative | 1,003 (94.6) | 57 (5.4) | Ref | 0.27 |
| Positive | 555 (95.9) | 24 (4.2) | 0.8 (0.5-1.2) |  |
| **HIV status** |  |  |  |  |
| Negative | 1,782 (95.6) | 82 (4.4) | Ref | 0.15 |
| Positive | 399 (94.1) | 25 (5.9) | 1.4 (0.9-2.2) |  |
| Unknown | 106 (93.8) | 7 (6.2) | 1.4 (0.6-3.2) |  |
| **Self-reported antiretroviral therapy (ART) use among HIV-positive** |  |  |  |  |
| No | 35 (94.6) | 2 (5.4) | Ref | 0.89 |
| Yes | 364 (94.1) | 23 (5.9) | 1.1 (0.3-4.9) |  |
| ***Trichomonas (T.) vaginalis* status (Ntot=1,556)** |  |  |  |  |
| Negative | 1,927 (95.4) | 92 (4.6) | Ref | 0.25 |
| Positive | 225 (93.8) | 15 (6.2) | 1.4 (0.8-2.5) |  |
| ***S. haematobium* by Circulating Anodic Antigen (CAA) (Ntot=1,685)** |  |  |  |  |
| Negative | 1,924 (95.4) | 93 (4.6) | Ref | 0.42 |
| Positive | 348 (94.3) | 21 (5.7) | 1.2 (0.8-2.0) |  |
| ***S. haematobium* by urine microscopy (Ntot=1,691)** |  |  |  |  |
| Negative | 2,168 (95.3) | 106 (4.7) | Ref | 0.40 |
| Positive | 119 (93.7) | 8 (6.3) | 1.4 (0.7-2.9) |  |

Percentages (%) on the second and third columns are calculated within each row total.
<sup>1</sup> *Molecular FGS* refers to PCR diagnosis of either home-based self-collected cervicovaginal swabs or clinic-based provider-collected cervicovaginal swabs.
<sup>2</sup> Visual FGS defined as a positive if confirmed by both reviewers 1 and 2, or in case of disagreement, by reviewer 3. The association between the single *visual FGS* and infection with HPV16 are presented in S5 Table
Abbreviations: AOR = Adjusted Odds Ratio, CI = Confidence Interval

## Discussion

To our knowledge, the *Zipime-Weka-Schista* study is the first to investigate the association between HR-HPV and FGS, diagnosed using both *visual* and *molecular* methods, in a large cohort of women with and without HIV in Zambia (27). We found a significant association between women with FGS defined as DNA detected by qPCR from cervicovaginal swabs (*molecular FGS*) and HPV16/18/45, the most frequently associated with cervical precancer and cancer. In contrast, no significant association was found between *visual FGS,* diagnosed using colposcopy, and HR-HPV infection. These findings highlight the need to recognize FGS as a sexual and reproductive health (SRH) condition and to integrate FGS screening into cervical cancer control programmes to improve early detection and reduce the dual burden of diseases in *S. haematobium* endemic settings (7).

Women with *molecular FGS* were almost 1.7 times more likely to test positive for HPV16/18/45, the most oncogenic genotypes which contribute to the majority of cervical precancer and cancer globally (12,13). To our knowledge, this is the first study to compare *molecular FGS* with molecular detected HR-HPV infection from cervicovaginal self-swabs for both infections. This finding suggests that FGS may play a role in the natural history of cervical HPV infection. HPV infects the undifferentiated, proliferating basal cells of the cervicovaginal epithelium through microabrasion, often resulting from co-existence of other STIs or co-factors that compromise the epithelial barrier (31). The epithelial damage of the anogenital mucosa caused by *S. haematobium* egg deposition in women with FGS may facilitate HPV viral entry (18). Further, the chronic genital inflammation caused by the *S. haematobium* egg deposition in the female genital tract may impair local immune responses, promoting immune evasion and HR-HPV persistence (18,31). Although the cross-sectional nature of this analysis does not allow to distinguish between incident and persistent HR-HPV infections, the observed association between *molecular FGS* and HPV16/18/45 support the hypothesis that FGS may contribute to HPV persistence. This aligns with findings from a recent study in Tanzania that followed 96 HIV-negative women and found higher odds of HR-HPV persistence at 9-12 months among women with *Schistosoma* infection, diagnosed by CAA (32). This analysis did not assess for genital involvement of schistosome infection but it supports the hypothesis that the schistosomiasis-related inflammation may contribute to persistence of HR-HPV infection (32). Longitudinal data from the ongoing *Zipime-Weka-Schista* study will further evaluate whether FGS promotes persistence or delays clearance of HR-HPV (27).

Women with egg-patent urinary *S. haematobium* infection were more likely to have prevalent HR-HPV infection, compared to those without. In contrast, no significant association was observed between schistosomiasis detected by CAA and HR-HPV infection. This may reflect the differences between diagnostic method (30). CAA is a highly sensitive diagnostic method for active *Schistosoma* infection, detecting antigens released by live adult worms in the host’s bloodstream (28). However, it does not provide information on egg deposition or differentiate between species (28). The absence of an observed association between CAA and HR-HPV but a positive association with egg-patent urinary schistosomiasis highlights the complexity of active infection in the pathogenesis of HR-HPV in the presence of *S.haematobium*.

In our study, *visual FGS,* diagnosed by colposcopy, was not associated with HR-HPV prevalence. Previous studies on this association reported mixed results, likely due to differences in outcome definitions, with one study including any HPV type (both high-and low-risk genotypes) and the other any HR-HPV type (24,25). Although colposcopy is widely used to assess FGS-related morbidity, its scalability in *S. haematobium* endemic settings is limited by high cost, infrastructure requirements, and need for trained personnel (2,6). Importantly, the mucosal changes in *visual FGS* positive women can be similar to changes from other SRH, including cervical lesions caused by HPV, resulting in limited diagnostic specificity (6,7). In our study, agreement between two expert reviewers in detecting *visual FGS* was low, consistent with previous research in Zambia (6). This reinforces the limitation of visual diagnosis for FGS and highlights the need for standardised, scalable diagnostic methods (6). Emerging technologies such as computer vision, powered by artificial intelligence (AI), are being explored to aid in the visual diagnosis of FGS (33,34,35). Computer vision tools are in the later stages of validation for the automated detection of cervical precancer and cancer in colposcopy images, potentially offering a point of integration for visual diagnosis of both FGS and cervical cancer (36).

Prevalence of HR-HPV was highest among women young women 15-25 years old, consistent with previous studies (37–40). Across our study population, prevalence of *molecular FGS* decreased with increasing age and peaked among adolescents aged 15-19 years old. In contrast, more older women (aged 41-50 years old) were positive for *visual FGS*. These findings are consistent with previous studies (3, 4, 29, 42). The similar age distribution supports possible opportunities to integrate FGS into cervical cancer control programmes (7). This is particularly timely as Zambia transitions from VIA to HPV DNA-based cervical screening programmes (20). For instance, existing self-collection platform and laboratory infrastructures for cervical cancer screening could include molecular FGS testing in endemic districts, improving earlier detection and more efficient use of resources (7). Younger women are more susceptible to HR-HPV infection due to behavioural and biological factors, including early sexual debut, multiple sexual partners, low contraception use, and cervical ectopy (37–40). In *S. haematobium* endemic settings, women in this age group also tend to have higher intensity of *S. haematobium* infection and higher rates of *Schistosoma* DNA retrieval from the genital tract, likely due to increased vascularization which promotes inflammation and parasite DNA shedding on the mucosal surface (3,29,42). In contrast, older women typically show higher prevalence of visually diagnosed lesions from long-standing *S. haematobium* egg deposition or chronic infection, reflecting the cumulative burden of FGS over time (3,29,42). This age-specific pattern suggests that, in *S. haematobium* endemic settings, *molecular FGS* screening could be integrated with HPV vaccination and education programmes targetting younger girls (aged 9-14 years old), while community-based integrated HPV and *molecular FGS* screening can be introduced for women of reproductive age eligible for cervical cancer screening (age 25 or 30 depending on HIV status) (19,20). For older women (aged over 35 years), FGS screening can be integrated into existing cervical cancer clinic-based screening and/or treatment programmes (19,20). These targeted age-specific efforts offer an approach for addressing the dual burden FGS and cervical cancer vulnerable populations.

Prevalence of any HR-HPV did not vary by HIV status, differing from previous studies showing higher HR-HPV prevalence and persistence among women living with HIV (15). This finding may reflect the high proportion of participants on ART and younger age in our study population. ART has been reported to reduce HPV persistence and precancer and cancer incidence, especially when initiated early following HIV acquisition and before significant immune suppression (17). Yet, these benefits have primarily been demostranted in monitored cohorts where women achieve sustained viral suppression through long-term ART adherence and regular cervical cancer screening. In our community-based study, ART status was self-reported, and baseline data on CD4+ T-cell count and HIV plasma viral load were not available at the time of this analysis, limiting our ability to assess immune control and its associations with HPV infection. Of note, we observed a 1.7-fold higher prevalence of the most oncogenic HPV types 16/18/45 among women living with HIV compared to HIV negative women, consistent with the literature (43,44,45).

With over 2,500 women recruited in the cohort, this is the largest study to date to evaluate the association between HR-HPV prevalence and FGS, diagnosed using both molecular and visual methods. The integrated nature of the study design, combining home and clinic-based sampling with multiple diagnostic methods allowed for comprehensive screening for FGS and SRH co-infections. The feasibility of this approach is reflected in the succesful recruitment and diagnostic completeness obtained. There are some limitations worth noting. Histological examination of cervical tissues for the detection of *Schistosoma* eggs was not performed, limiting the ability to compare *molecular* and *visual FGS* diagnoses against a reference standard, as both have imperfect sensitivity and specificity (2,6,46). The reliability of *visual FGS* diagnosis was limited by low agreement between two expert reviewers, highlighting the need for standardised visual diagnostic methods. Implementation of molecular FGS testing in routine clinical or programmatic setting is hindered by cost, technical requirements, and the need for trained personnel. Moreover, variations in assay design may affect comparability across laboratories for PCR testing. Although this study accounted for HIV and *T. vaginalis* status, it did not address potential confounding by other STIs, including *Chlamydia trachomatis, Neisseria gonorrhoea, Mycoplasma genitalium, Herpes simplex* virus, syphilis, and bacterial vaginosis, all of which can affect cervical inflammation and epithelial integrity (47). HIV status was self-reported for most participants which may have introduced reporting bias. Data on CD4+ T-cell count, time on ART, ART adherence and HIV plasma viral load among the study participants was not collected, potentially introducing further confounding. Further, the cross-sectional design of this analysis does not allow to distinguish between incident and persistent HR-HPV infections, limiting causal inferences on the association between FGS and HR-HPV. Longitudinal data from the ongoing *Zipime-Weka-Schista* study (27) will address some of these limitations in future analyses.

This study contributes new evidence on the intersection of FGS, HR-HPV and SRH among women of reproductive age living, highlighting a significant association between molecular FGS and the most oncogenic HPV genotypes (HPV16/18/45). In line with the Sustainable Development Goals and the WHO targets to eliminate cervical cancer and schistosomiasis, our findings support the need for integrated strategies using highly specific, field-deployable, and cost-effective diagnostic methods.

## Data Availability

Due to the sensitive nature of the data collected in the Zipime-Weka-Schista, data will be available upon request.

## Acknowledgements

We would like to thank and acknowledge all the study participants and the communities in Kafue and Livingstone for their engagement and support. We acknowledge the invaluable contribution of Zipime-Weka-Schista community workers, field teams and midwives. We also thank the wider Zipime-Weka-Schista Study team who contributed towards the design and implementation of the study and Isaac Mshanga for his work on community sensitization and mobilization. We are grateful for our partners in Zambia, including the Ministry of Health and District Health Management Teams, as well as the administrative support teams at both Zambart and LSHTM. We gratefully acknowledge the team at the Leiden University Medical Centre (LUMC) for performing the CAA analysis. We thank the ODK forum for assistance with questionnaire construction and Chrissy Roberts (LSHTM) for his invaluable support with 3D printing of teaching models.

