## Supplementary material for "Association between female genital schistosomiasis and high-risk human papillomavirus among women of reproductive age in Zambia: the Schista study"

**Supplementary material for "Cross-sectional association between Female Genital Schistosomiasis and High-risk Human Papillomavirus across three communities in Zambia: The Schista study"**

**Supplementary Text:**

### S1 Text: Sample transportation and storage

Samples collected both at home and in clinic were transported on the same day in cool boxes to the field laboratory for further storage and further processing. The cervicovaginal swabs were stored at -80°C after arrival at the field laboratory. Cervicovaginal swabs were then transported under frozen to Zambart central laboratories (Lusaka, Zambia) for further DNA extraction and analysis. Cervicovaginal lavages (CVLs) were stored at -20°C at Zambart central laboratories (Lusaka, Zambia) and will be used for future analysis. U A sample of 2mL of urine per participant was stored at 4°C in the field laboratory and shipped to Zambart central laboratories (Lusaka, Zambia) and then to Leiden University Medical Centre.

### S2 Text: Egg-patent urinary *S. haematobium*

A single 10mL urine sample was used to detect *S. haematobium* egg-patent infection by microscopy. The analysis was performed at the field laboratory on the same day as sample collection. Urine samples were centrifugated in aliquots and examined by microscopy by a trained technician for the detection of *S. haematobium* eggs. Women with evidence of urinary *S. haematobium* were treated free of charge with 40mg/kg praziquantel (27).

### S3 Text: Circulating Anodic Antigen (CAA)

A 2mL urine sample was shipped to Leiden University Medical Centre (LUMC), The Netherlands for *Schistosoma* CAA detection. Living schistosome worms excrete CAA into the host's bloodstream and CAA-levels are a reflection of the worm burden (1–3). A quantitative lateral flow assay utilizing up-converting reporter particles was used to detect CAA in urine samples. For this assay, a CAA value indicating levels >0.6 pg/mL (lower limit of detection) was considered positive (1–3).

### S4 Text: PCR Detection of *Schistosoma* DNA from cervicovaginal swabs

PCR of cervicovaginal swabs collected either at home or in clinic was performed at Zambart central laboratories (Lusaka, Zambia). Total nucleic acid (TNA) extraction (PrimeXtract™ Longhorn Vaccines & Diagnostics, USA) was performed as per manufacturer's instructions, using a simplified silica-based spin column extraction process. A 500µL PrimeStore® MTM

((PS-MTM) sample was used per extraction column, with final elution in 50µL manufacturer provided elution buffer. Extracted nucleic acid was stored at -20◦C until use.

qPCR was performed for the amplification and detection of a *Schistosoma*-specific 77bp fragment of ITS-2 (Internal Transcribed Spacer), as previously reported with some minor adaptation (4,5) (3,29). Briefly, 5µL TNA was used in each reaction. HotStar Taq Polymerase Master Mix (Qiagen) was used for each reaction which also contained 3.5mM $MgCl_2$; 0.1mg/ml bovine serum albumin (BSA); ROX reference dye (Invitrogen); custom fluorophore (FAM) labelled probe (TIB MolBiol Germany; Ssp78T_FAM: 5′-TGGGTTGTGCTCGAGTCGTGGC; quencher: BHQ1) and forward/reverse primers (Ssp48F: 5′-GGTCTAGATGACTTGATYGAGATGCT and Ssp124R: 5′-TCCCGAGCGYGTATAATGTCATTA). Thermal cycling conditions were done as follow: 15min at95◦C; followed by 50 cycles of 95◦C for 15 seconds; 60◦C for 30 seconds; 72◦C for 30 seconds. qPCR was performed on a StepOnePlus (Applied Biosystems; Thermofisher) thermal cycler or CFX96 (BioRad) thermal cycler, according to manufacturer instructions.

Results were reported in cycle threshold values (Ct-values). Ct values were generated in reference to a standard curve, derived from a series of known dilutions (serial 1:5) of TNA isolated from *S. haematobium eggs*, extracted as above, and quantified using a Qubit™ 4 Fluorometer (Thermofisher).

**S5 Text: GeneXpert HPV testing method**

DNA extraction for self-collected cervicovaginal swabs stored in PrimeStore MTM was performed using a modified PrimeXtract™ spin column protocol. In this modified procedure, instead of the standard 500 µL input volume, 700 µL of PS-MTM was applied per column. The extraction included sequential washes with Wash 1 and Wash 2 buffers. A double elution step was introduced in place of the standard single elution, using the manufacturer-supplied elution buffer, resulting in a final eluate volume of 100 µL.

Each GeneXpert HPV cartridge included a sample adequacy control (SAC) and a Probe Check Control (PCC). SAC reagent detects the single copy human gene (*Hydroxymethylbilane synthase (HMBS))* to confirm adequate human cellular material for valid HPV detection. The PCC verifies reagent rehydration, PCR tube filling in the cartridge, probe integrity, and dye stability

### S6 Text: Interpretation of GeneXpert HPV Results

In addition to possible positive and negative outcomes, there are three other possible outcomes:

1. Invalid
   Presence or absence of HPV target DNA cannot be determined and repeat the test according to the instructions required.
   Criteria for Invalids:
   - SAC: FAIL meaning that the Ct is not within the valid range and/or a fluorescence endpoint below the threshold setting.
   - PCC: PASS; all probe check results pass.

2. Error: Presence or absence of HPV target DNA cannot be determined and repeat the test according to the instructions required.
   Criteria for errors:
   - SAC: No Result
   - PCC: Fail. If the probe check passed, the error is caused by the maximum pressure limit exceeding the acceptable range or by a system component failure.

3. No Result: Presence or absence of HPV target DNA cannot be determined, repeat test according to the instructions required. A NO RESULT indicates that insufficient data were collected. For example, the operator stopped a test that was in progress, or a power failure occurred.
   Criteria:
   - HPV: NO RESULT
   - SAC: NO RESULT
   - PCC: Not Applicable

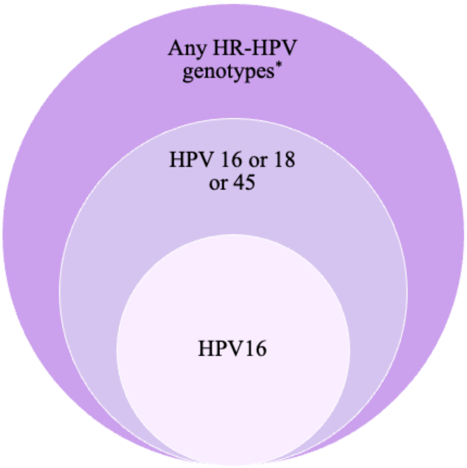


*Positive for any of HPV 16, 18, 45, 31, 33, 35, 52, 58, 51, 59, 39, 56, 66, 68

**Figure S1:** High-risk (HR-) human papillomavirus (HPV) categories

**Supplementary results**

**Self-reported sexual and reproductive health behaviours and outcomes**

Women presented with a range of sexual and reproductive (SRH) symptoms (Figure S1). SRH symptoms ranged from 3.8% for intermenstrual bleeding to 29.7% for vaginal itching. Women with *any* HR-HPV infection more frequently reported postcoital bleeding (HR-HPV positive 10.1% vs HR-HPV negative 6.7%; Chi2 p-value=0.004), dyspareunia (HR-HPV positive 17.1 % vs HR-HPV negative 13.7%; Chi2 p-value=0.04), intermenstrual bleeding (HR-HPV positive 5.1% vs HR-HPV negative 3.2%; Chi2 p-value=0.02), and genital sore (HR-HPV positive 11.0% vs HR-HPV negative 8.4% Chi2 p-value=0.04) compared to those without (Figure S1).

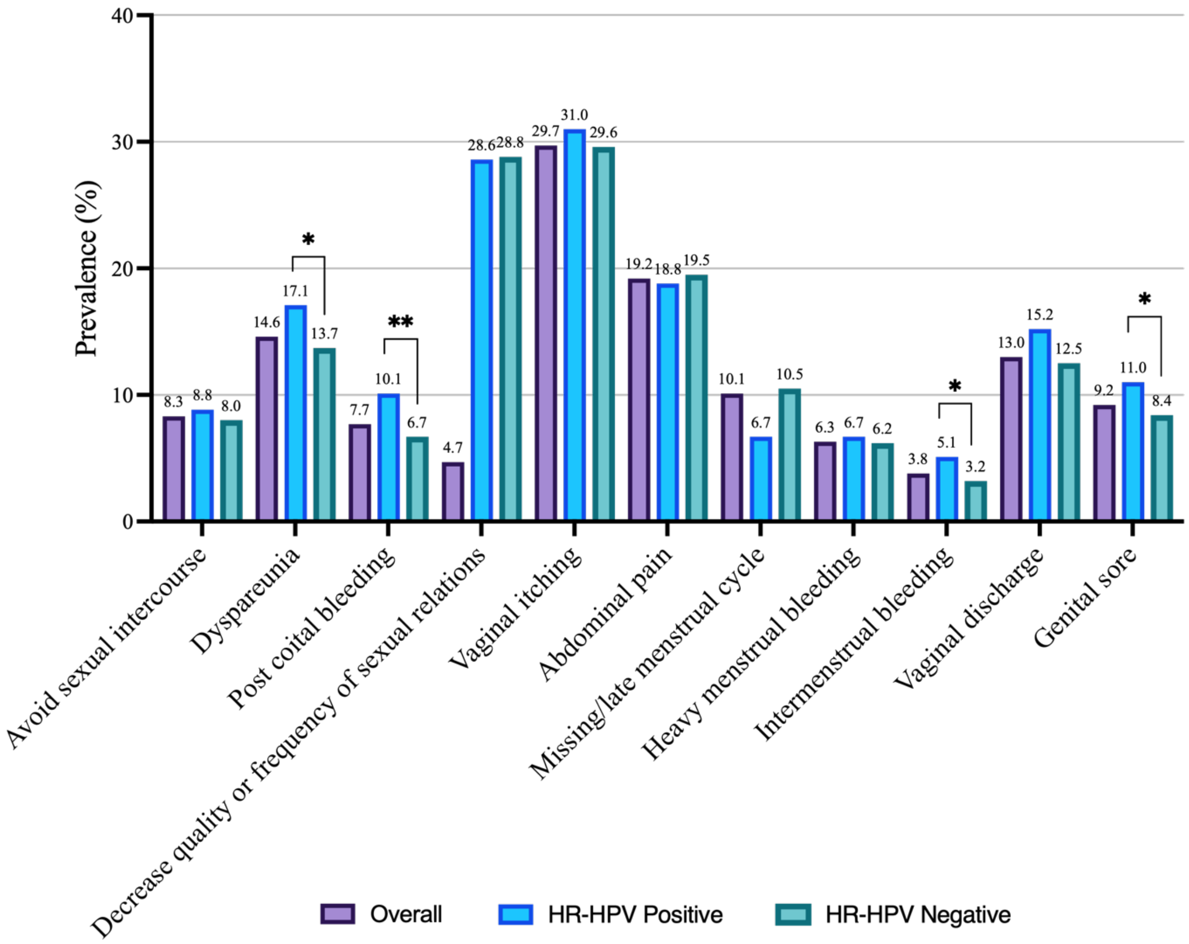


Note: * is for p<0.05, ** p<0.01. Asterisks indicate statistically significant differences in symptoms prevalence between HR-HPV positive and HR-HPV negative individuals.

**S2 Figure:** Prevalence of sexual and reproductive signs and symptoms by HR-HPV infection.

**S1 Table:** Positive diagnostic test results by age categories.

| Age categories | Molecular FGS N (%) | Visual FGS N (%) | Urine CAA N (%) | Urine microscopy N (%) | Any HR-HPV type N (%) | HPV16/18/45 N (%) | HPV16 only N (%) |
|---|---|---|---|---|---|---|---|
| **15-19** | 48 (14.6) | 63 (31.0) | 70 (21.4) | 41 (12.4) | 112 (36.0) | 32 (10.3) | 15 (4.8) |
| **20-25** | 53 (7.9) | 120 (27.9) | 98 (14.6) | 35 (5.2) | 223 (35.1) | 66 (10.4) | 33 (5.2) |
| **26-30** | 26 (4.7) | 134 (34.9) | 84 (15.6) | 18 (3.3) | 147 (27.9) | 55 (10.4) | 23 (4.4) |
| **31-35** | 13 (3.8) | 72 (32.3) | 49 (14.4) | 14 (4.1) | 78 (24.2) | 32 (9.9) | 21 (6.5) |
| **36-40** | 11 (3.8) | 89 (42.2) | 37 (12.8) | 10 (3.4) | 55 (19.8) | 18 (6.5) | 7 (2.5) |
| **41-45** | 7 (3.5) | 62 (45.3) | 32 (16.0) | 9 (4.5) | 47 (24.6) | 21 (11.0) | 9 (4.7) |
| **46-50** | 7 (5.0) | 55 (53.4) | 18 (12.8) | 5 (3.6) | 28 (20.6) | 7 (5.2) | 6 (4.4) |
| **Positives** | 165 | 595 | 388 | 132 | 690 | 231 | 114 |
| **Total** | 2,532 | 1,691 | 2,517 | 2,532 | 2,401 | 2,401 | 2,401 |
| **P-value*** | <0.001 | <0.001 | 0.05 | <0.001 | <0.001 | 0.26 | 0.45 |

*Chi-squared p-value for difference between groups

**Abbreviations:** CAA- circulating Anodic Antigen, FGS- Female Genital Schistosomiasis, HR-HPV – high-risk human papillomavirus, HPV-human papillomavirus

**S2 Table**: Association between individual *visual FGS* cervicovaginal lesions and presence of any HR-HPV

| | | | Univariable model | |
|---|---|---|---|---|
| | **HR-HPV negative** | **HR-HPV positive** | **Crude OR (95% CI)** | **P-value** |
| **Homogeneous yellow sandy patches** | | | | |
| Negative | 822 (69.2%) | 366 (30.8%) | Ref | 0.28 |
| Positive | 294 (72.1%) | 114 (27.9%) | 0.9 (0.7-1.1) | |
| **Grainy sandy patches** | | | | |
| Negative | 966 (69.9%) | 417 (30.2%) | Ref | 0.60 |
| Positive | 83 (72.2%) | 32 (27.8%) | 0.9 (0.6-1.4) | |
| **Abnormal blood vessels** | | | | |
| Negative | 1.005 (69.6%) | 439 (30.4%) | Ref | 0.05 |
| Positive | 92 (78.6%) | 25 (21.4%) | 0.6 (0.4-1.0) | |
| **Rubbery papules** | | | | |
| Negative | 1,045 (69.9%) | 451 (30.2%) | Ref | 0.82 |
| Positive | 3 (75.0%) | 1 (25.0%) | 0.8 (0.08-7.5) | |

**S3 Table:** Multivariable logistic regression model parameter for the association between *S. haematobium* infection by urine microscopy and *any* HR-HPV infection (N=2,390)

| **Variables** | | **Multivariable model** | |
|---|---|---|---|
| | | **AOR (95% CI)[1]** | **p-value** |
| **Urinary *S. haematobium* infection by microscopy** | Negative | Ref | 0.009 |
| | Positive | 1.7 (1.1-2.4) | |
| **Age (years)** | 15-19 | 1.7 (1.0-2.8) | <0.001 |
| | 20-25 | 2.0 (1.2-3.2) | |
| | 26-30 | 1.6 (1.0-2.5) | |
| | 31-35 | 1.4 (0.8-2.3) | |
| | 36-40 | 1.0 (0.6-1.8) | |
| | 41-45 | 1.4 (0.8-2.3) | |
| | 46-50 | Ref | |
| **Marital status** | Married | Ref | 0.08 |
| | Single | 1.7 (1.4-2.1) | |
| | Divorced | 1.4 (0.9-2.2) | |
| | Widowed | 2.4 (1.4-4.3) | |

[1]AOR = Adjusted Odds Ratio, CI = Confidence Interval
Model adjusted for FGS status, age, and marital status.
Confounding variables were selected based on a review of the literature and statistical significance from the univariable association with the outcome.

**S4 Table:** Association between individual *visual FGS* cervicovaginal lesions and presence of HPV16/18/45

| | | | Univariable model | |
|---|---|---|---|---|
| | **HR-HPV negative** | **HR-HPV positive** | **Crude OR (95% CI)** | **P-value** |
| **Homogeneous yellow sandy patches** | | | | |
| Negative | 1,062 (89.4%) | 126 (10.6%) | Ref | 0.19 |
| Positive | 374 (91.7%) | 34 (8.3%) | 0.8 (0.5-1.1) | |
| **Grainy sandy patches** | | | | |
| Negative | 1,242 (89.8%) | 141 (10.2%) | Ref | 0.61 |
| Positive | 105 (91.3%) | 10 (8.7%) | 0.8 (0.4-1.6) | |
| **Abnormal blood vessels** | | | | |
| Negative | 1,304 (90.3%) | 140 (9.7%) | Ref | 0.92 |
| Positive | 106 (90.6%) | 11 (9.4%) | 0.9 (0.5-1.8) | |
| **Rubbery papules** | | | | |
| Negative | 1,345 (89.9%) | 151 (10.1%) | Not enough data | |
| Positive | 4 (100.0%) | 0 | | |

**S5 Table**: Association between individual *visual FGS* cervicovaginal lesions and presence of HPV16 only

| | | | Univariable model | |
|---|---|---|---|---|
| | **HR-HPV negative** | **HR-HPV positive** | **Crude OR (95% CI)** | **P-value** |
| **Homogeneous yellow sandy patches** | | | | |
| Negative | 1,126 (94.8%) | 62 (5.2%) | Ref | 0.40 |
| Positive | 391 (95.8%) | 17 (4.2) | 0.8 (0.5-1.4) | |
| **Grainy sandy patches** | | | | |
| Negative | 1,313 (94.9%) | 70 (5.1%) | Ref | 0.45 |
| Positive | 111 (96.5%) | 4 (3.5%) | 0.7 (0.2-1.9) | |
| **Abnormal blood vessels** | | | | |
| Negative | 1,377 (95.4%) | 67 (4.6%) | Ref | 0.81 |
| Positive | 111 (94.9%) | 6 (5.1%) | 1.1 (0.5-2.6) | |
| **Rubbery papules** | | | | |
| Negative | 1,421 (95.0%) | 75 (5.0%) | Not enough data | |
| Positive | 4 (100.0%) | 0 | | |

**S6 Table :** Agreement between expert reviewers for the presence or absence of *visual FGS*

| | Reviewer 1 | | | |
|---|---|---|---|---|
| **Reviewer 2** | Visual FGS not detected | Visual FGS detected | Not evaluated | Total |
| Visual FGS not detected | 421 | 1,004 | 4 | 1,429 |
| Visual FGS detected | 30 | 254 | 1 | 285 |
| Not evaluated | 4 | 0 | 814 | 818 |
| Total | 455 | 1,258 | 819 | 2,532 |
